# Mixed-Effects Machine Learning improved prediction of Obstetric Anal Sphincter Injury: a nationwide multicenter cohort study

**DOI:** 10.64898/2026.09.29.26364280

**Authors:** Abdelhak Chahid Mohamed, Anita CJ Ravelli, JanWillem de Leeuw, Jeroen van Bavel, Ameen Abu-Hanna

**Affiliations:** Department of Medical Informatics, Amsterdam UMC, location University of Amsterdam, The Netherlands; Public Health Research Institute, Amsterdam, The Netherlands; Department of Obstetrics and Gynaecology, Ikazia Hospital, The Netherlands; Department of Obstetrics and Gynaecology, Amphia Hospital Breda, The Netherlands

**Keywords:** Obstetric anal sphincter injury, machine learning, mixed-effects modeling, artificial intelligence, LASSO variable selection, risk prediction, temporal validation

## Abstract

Existing prediction models for obstetric anal sphincter injuries (OASIS) often rely on traditional regression, lack external validation, and, when fitted on multicenter data, fail to account for hospital-level clustering. We aimed to develop and temporally validate mixed-effects machine learning models for OASIS and compare them with traditional regression models. A secondary aim was to evaluate whether macrosomia (≥4000 g) could substitute for continuous birth weight. We conducted a retrospective nationwide cohort study of singleton, term (≥37 weeks), cephalic vaginal deliveries in the Dutch Perinatal Registry. Data from 2016 to 2019 were used for model development with three-fold cross-validation; data from 2020 were reserved for temporal validation. Candidate predictors included 18 demographic, obstetric, and neonatal factors. We developed six models: mixed-effects gradient boosting (MEGB), mixed-effects LASSO (ME-LASSO), and backward stepwise logistic regression, each with either birth weight or macrosomia. Discrimination was assessed by the area under the receiver operating characteristic curve (AUC), calibration by calibration plots, and prediction accuracy by the Brier score. Among 614,255 deliveries, OASIS occurred in 2.6%. In temporal validation including macrosomia, MEGB achieved the highest discrimination (AUC 0.727; 95% CI 0.719–0.735), significantly outperforming ME-LASSO (AUC 0.719; 95% CI 0.710-0.727; P<.0001) and stepwise logistic regression (AUC 0.714; 95% CI 0.706–0.722; P<.0001). Including birth weight marginally improved performance, with macrosomia serving as an effective proxy. Consistently important predictors were nulliparity, prolonged second stage, mediolateral episiotomy, advanced maternal age, operative vaginal delivery, macrosomia, and previous cesarean delivery. Uncalibrated MEGB showed poorer calibration, but isotonic recalibration substantially improved it. After calibration, all models showed excellent agreement between predicted and observed risks. MEGB provided improved discrimination and comparable calibration relative to ME-LASSO and stepwise logistic regression. Replacing continuous birth weight with a binary macrosomia indicator resulted in minimal performance loss, suggesting it may suffice for clinical risk prediction across all three modeling approaches.

**Author summary:** Obstetric Anal Sphincter Injuries (OASIS) after giving birth, can lead to long-term issues such as incontinence, pelvic floor dysfunction, chronic pain, and sexual dysfunction. Identifying women at increased risk before birth could support counselling and help clinicians take preventive measures. Although several prediction tools have been developed, many do not account for possible differences between hospitals, where clinical practices and rates of OASIS may vary.

Using nationwide data from more than 600,000 births in the Netherlands, we developed prediction models for OASIS that also accounted for differences between hospitals. We used machine-learning approaches and compared their performance with that of a traditional regression model. The machine-learning models performed slightly better in predicting OASIS. We also found that classifying babies as large (birth weight above 4,000 g) performed almost as well as using exact birth weight. This is relevant because fetal weight can only be estimated before birth using ultrasound. After further external validation, these models may support earlier and more individualized assessment of OASIS risk.

## Introduction

Obstetric anal sphincter injuries (OASIS) affect 2–4% of vaginal deliveries [1–4] and represent a serious complication, with long-term sequelae including fecal incontinence, pelvic floor dysfunction, chronic pain, and sexual dysfunction [5].

Prediction of OASIS remains challenging due to its multifactorial etiology and the complex interplay of risk factors. Nonetheless, accurate prediction is crucial, as it enables clinicians to effectively inform and counsel pregnant women. Several maternal, fetal, and delivery-related factors have been associated with increased OASIS risk. Nulliparity, prolonged labor, instrumental delivery, previous cesarean delivery and increased birth weight are consistently reported as significant contributors [4,6]. OASIS risk also varies by ethnic background, with several studies reporting higher rates among women of South Asian ethnicity compared with European women [3,7,8]. While mediolateral episiotomy during operative vaginal delivery may reduce OASIS risk [8,9], it also carries potential harms.

A recent systematic review described that, 25 prediction models for OASIS have been developed, with reported discrimination ranging from 0.64 to 0.83 [10]. However, that systematic review found that all had methodological limitations, including small or poorly generalizable cohorts and risk of bias, and that none had undergone external geographical validation, so even the higher discrimination values may be optimistic. Key recurring limitations concern sample size, variable selection, model specification, accounting for the nested structure of data in hospitals or regions, and external validity. Patient characteristics and clinical practices are nested within hospitals or regions, creating a hierarchical structure where observations from the same region resemble each other more than those from different regions. Clinical practices (e.g., episiotomy use, labor management) often vary systematically by hospital or by region.

Traditional models assume independent observations, which, when violated by clustering, can bias estimates and standard errors. Mixed-effects models are better suited for this context as they account for fixed effects, predictors that influence OASIS risk across the population, and random effects that capture region-specific variation. This approach addresses intra-cluster correlation and yields more reliable, generalizable estimates [11,12].

In addition, most existing OASIS prediction models rely on traditional multivariable regression [2,3,13–15], which may not capture complex interactions or nonlinear relationships among risk factors. These models are frequently developed from single-center cohorts and depend on retrospective variables such as actual birth weight, which is only available post-delivery and limits real-time applicability [16,17]. While ultrasound can estimate fetal weight, its accuracy is variable[18,19], and using estimated fetal weight (EFW) as a continuous predictor may introduce additional noise. Instead, converting EFW into a binary variable such as macrosomia, provides a more practical and interpretable approach, reduces sensitivity to measurement error, and better supports real-time clinical decision-making.

To address these limitations, we applied mixed-effects gradient boosting (MEGB) and mixed-effects LASSO (ME-LASSO), comparing them to baseline logistic regression and validating them temporally. We developed two versions of each model: one using continuous birth weight and one using macrosomia as a binary proxy, allowing assessment of whether antenatally estimable macrosomia can substitute for exact birth weight for pre-delivery risk stratification.

Our aim is twofold: First, to evaluate the performance of modern machine learning methods, specifically mixed-effects gradient boosting (MEGB) and mixed-effects penalized logistic regression (ME-LASSO), compared to stepwise logistic regression for predicting OASIS among nulliparous and multiparous women with term vaginal deliveries. Second, to evaluate whether antenatally estimable macrosomia can serve as a proxy for birth weight, supporting pre-delivery OASIS risk stratification.

## Material and methods

### Ethics Statement

The Perined Ethics Committee approved the use of registry data for this study (approval number 17.18). Data were collected anonymously by Perined, the Netherlands Perinatal Registry. Given the non-invasive, retrospective design of this registry-based cohort study, no additional ethical approval or individual informed consent was required under Dutch law.

### Study Design and Population

This retrospective cohort study used anonymized data from the Dutch Perinatal Registry (www.perined.nl), covering over 96% of deliveries nationwide [20]. The PERINED dataset includes pregnancies from 22 weeks’ gestation onward, with detailed maternal, labor, delivery, and neonatal information.

We included women with singleton, term (≥37.0–42.0 weeks) vaginal deliveries in cephalic presentation between January 1, 2016, and January 1, 2021. Both nulliparous and multiparous women were included, from all nine healthcare regions.

### Outcome and Predictors

The primary outcome, obstetric anal sphincter injury (OASI), was defined as any third-degree (3a, 3b, 3c) or fourth-degree perineal tear, in accordance with international classification standards [21], registered as subtotal or total perineal ruptures, respectively. Tears were diagnosed and classified by the attending midwife, obstetrician, or other perinatal healthcare provider immediately after delivery, as part of routine clinical registration.

Candidate predictors were selected based on clinical relevance, literature, and availability in the national perinatal registry. These included maternal demographics (age, parity, ethnicity), obstetric history (prior cesarean), pregnancy-related conditions (hypertension, diabetes), labor characteristics (fetal head presentation, induction, epidural analgesia, gestational age, mode of delivery, duration of second stage, fetal distress), and neonatal factors (birth weight, macrosomia and fetal sex). Predictors requiring clinical judgment, such as fetal head presentation and fetal distress, were similarly assessed by the attending midwife or obstetrician as part of routine care; the qualifications or demographic characteristics of the specific assessor were not recorded in the registry.

### Data Processing

To improve numerical stability during model fitting, we standardized maternal age (years), gestational age (weeks), duration of the second stage of labor (minutes), and birth weight (grams). These transformations, performed prior to modeling, did not affect variable inclusion or interpretation but improved convergence.

Missing values were handed using multilevel predictive mean matching mice package in R, with hospital included as a clustering variable to account for the hierarchical data structure. Analyses were performed on a single completed dataset [22].

### Statistical analysis

Statistical analysis followed TRIPOD+AI guidelines for prediction modeling, and STROBE guidelines for observational study reporting. We developed six models: MEGB, ME-LASSO, and stepwise logistic regression, each with birth weight (Model I) and with macrosomia instead (Model II). Hyperparameters were tuned using three-fold cross-validation, as 5- to 10-fold was computationally prohibitive for MEGB and ME-LASSO, while the large sample size left sufficient data per fold for stable tuning. No separate sample size calculation was performed a priori, as all eligible registry cases were included, yielding a large analytic cohort (n = 614,255) with 16,182 OASI events. This event count substantially exceeds conventional minimum thresholds for the number of candidate predictors considered, and model complexity was further controlled through regularisation (ME-LASSO), hyperparameter tuning with cross-validation (MEGB), and backward variable selection (logistic regression). Together, the large sample size, high event count, and constraint on model complexity were considered sufficient to ensure stable and reliable model development. Analysis code is available at https://github.com/achahid/OASI-prediction-model.

#### Mixed Effects Machine Learning (MEGB)

We implemented the MEGB framework to jointly model nonlinear predictor effects and region-level clustering in OASI risk as described by Che Ngufor et al [23]. This EM-based approach alternates between estimating population-level patterns, via machine learning, and adjusting for region-level variability using a generalized linear mixed model (GLMM). Hyperparameters, including number of trees (150–250), tree depth (4–6), learning rate (0.05–0.1), and minimum observations per terminal node (15–25), were tuned via grid search with 3-fold cross-validation on the training set. The combination achieving the highest area under the ROC curve (AUC) was selected for the final model. MEGB’s classification threshold was optimized to jointly balance sensitivity and specificity, rather than using a fixed 0.5 cutoff.

Predictor importance was assessed using the relative influence metric from gradient boosting, which quantifies each variable’s contribution based on split frequency and loss reduction. Unlike regression models, gradient boosting does not yield interpretable coefficients; thus, variable importance provides a useful alternative for ranking predictors.

#### Mixed Effects LASSO (ME-LASSO)

We performed simultaneous selection of fixed and random effects using Regularized Penalized Quasi-Likelihood (PQL) as described by Hui et al [24]. This method applies adaptive LASSO penalties to shrink fixed-effect coefficients (e.g., episiotomy, birth weight) and group-penalize random effects (e.g., hospital/region-level intercepts), enabling efficient model fitting without computationally intensive integration.

The algorithm initializes the model parameters with PQL estimates and iteratively updates fixed effects with adaptive LASSO penalties while penalizing random effects covariance matrices via quasi-likelihood, continuing until coefficient changes stabilize. Hyperparameters for the shrinkage penalty (λ) were optimized using 3-fold cross-validation, selecting the value that maximized AUC.

For interpretability, variables selected by ME-LASSO were refitted in an unpenalized mixed-effects logistic regression to obtain unbiased odds ratios and confidence intervals, following Hui et al [24].

#### Stepwise Backward Logistic Regression

For comparison, we developed a multivariable logistic regression model using stepwise backward selection based on the Akaike Information Criterion AIC, without accounting for hospital clustering. Model performance was assessed by 3-fold cross-validation (AUC), and final coefficients, odds ratios, and 95% confidence intervals were reported.

### Model Performance

Performance was evaluated in the training and temporal validation datasets using discrimination (AUC), calibration (intercept, slope, Brier score), sensitivity, and specificity. Isotonic recalibration was applied on the development dataset to improve calibration before validation. Predictor contributions were assessed via variable importance (MEGB) and variable selection (ME-LASSO, backward stepwise logistic regression).

### Temporal Validation

To evaluate generalizability and robustness over time, we trained models on 2016–2019 data and temporally validated on 2020 data. This approach mimics real-world deployment by testing on future observations, detects overfitting and assessing robustness to changes in clinical practice or patient characteristics over time [25].

### Software

Statistical analysis was performed using R version 4.4.2.

#### Ethical Considerations

Ethical approval for this analysis of the pseudorandomized registry data was granted by the Perined Scientific Review Board (Approval #17.18). Under Dutch law, retrospective studies using anonymized registry data are exempt from additional ethics review.

## Results

### Study Population

Of 614,255 vaginal deliveries recorded between 2016 and 2020 in the Dutch Perinatal Registry, 262,315 were nulliparous (42.7%) and 351,940 were multiparous (57.3%) women with singleton, term (≥37 weeks), and cephalic live births, included after applying eligibility criteria (Fig 1).

**Fig 1.**
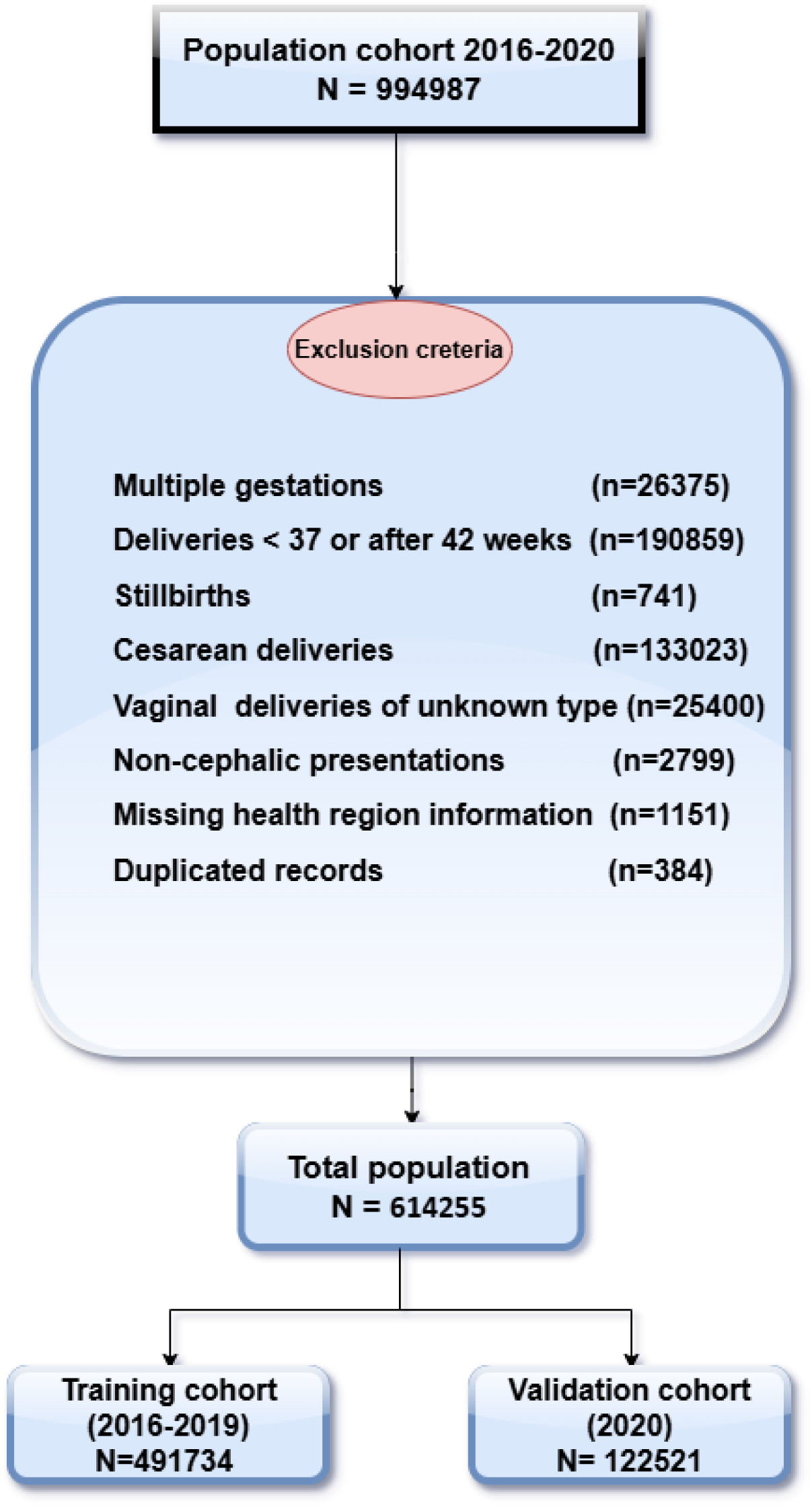
Flow diagram of the selected study population. Deliveries recorded in the Dutch Perinatal Registry between 2016 and 2020 were screened for eligibility. After exclusions detailed in the diagram, the final population was divided into a development cohort (2016–2019) and a temporal validation cohort (2020).

OASI occurred in 16,182 cases (2.6%). Missing values (duration of second stage, 6%; birth weight, 0.15%) were addressed using multilevel imputation.

### Baseline characteristics

Baseline characteristics for the 2016–2019 cohort are shown in Table 1. Compared to women without OASIS, those with OASIS had higher birth weight infants (3649 vs. 3495 grams) and experienced longer second stage of labor (44.5 vs. 31.9 minutes). Nulliparity (65% vs. 42%), prior cesarean delivery (5% vs. 3.5%), and macrosomia (22% vs. 13.7%) were more common among women with OASIS. The validation cohort was comparable to the development cohort (Table 1).

**Table 1.** Baseline characteristics of the women by OASIS status in the training and validation sets.

| Characteristics |  | Training :<br>year 2016- 2019 |  |  | Validation :<br>year 2020 |  |  |
| --- | --- | --- | --- | --- | --- | --- | --- |
|  |  | OASIS | NO-OASIS | Total | OASIS | NO-OASIS | Total |
| **Total n (%)** |  | 12,859<br>(2.6) | 478,875<br>(97.4) | 491,734 | 3323<br>(2.7) | 119,198<br>(97.3) | 122,521 |
| **Maternal age in years, mean, SD** |  | 30.3<br>(4.2) | 30.7 (4.6) | 30.6<br>(4.6) | 30.4<br>(4.25) | 30.9<br>(4.6) | 30.8<br>(4.6) |
| **Gestational age at delivery in completed weeks, n (%)** | 37 | 508<br>(3,9) | 32,465<br>(6.8) | 32,973<br>(6.7) | 130<br>(3.9) | 7747<br>(6.5) | 7877<br>(6.4) |
|  | 38 | 1502<br>(11,7) | 75,416<br>(15,8) | 76,918<br>(15.6) | 427<br>(12.8) | 19357<br>(16.2) | 19784<br>(16.1) |
|  | 39 | 3248<br>(25,3) | 123,741<br>(25,8) | 126,989<br>(26) | 840<br>(25.3) | 31009<br>(26.0) | 31849<br>(26.0) |
|  | 40 | 4548<br>(35.4) | 153,937<br>(32.1) | 158,485<br>(32) | 1167<br>(35.1) | 38369<br>(32.2) | 39536<br>(32.3) |
|  | 41 | 2823<br>(22.0) | 87,104<br>(18,2) | 89,927<br>(18.3) | 724<br>(21.8) | 21613<br>(18.1) | 22337<br>(18.2) |
|  | 42 | 230<br>(1.7) | 6,212<br>(1.3) | 6442<br>(1.3) | 35<br>(1.0) | 1103<br>(0.9) | 1138<br>(0.9) |
| **Ethnicity, n (%)** | Europea<br>n | 10,130<br>(79) | 376,075<br>(78) | 386,205<br>(78) | 2543<br>(76) | 91723<br>(77) | 94266<br>(77) |
|  | South<br>Asian | 263<br>(2.0) | 5,257<br>(1.0) | 5520<br>(1.1) | 89<br>(2.7) | 1403<br>(1.2) | 1492<br>(1.2) |
|  | Asian | 576<br>(4.5) | 15,486<br>(3.2) | 16062<br>(3.2) | 146<br>(4.4) | 4052<br>(3.4) | 4198<br>(3.4) |
|  | Other<br>non-Western | 1,890<br>(14.7) | 82,057<br>(17.1) | 83947<br>(17.1) | 545<br>(16.4) | 22020<br>(18.5) | 22565<br>(18.4) |
| **Fetal presentation, n (%)** | Occipito-anterior | 12102<br>(94.1) | 457,670<br>(95.6) | 469,772<br>(95.5) | 3127<br>(94.1) | 114069<br>(95.7) | 117196<br>(95.7) |
|  | Occipito-<br>posterior | 757<br>(5.9) | 21,205<br>(4.4) | 21962<br>(4.4) | 196<br>(5.9) | 5129<br>(4.3) | 5325<br>(4.3) |
| **Fetal birth weight in gram mean (SD)** |  | 3649<br>(456) | 3495<br>(461) | 3498<br>(462) | 3642<br>(446) | 3505<br>(460) | 3508<br>(460) |
| **Induction of labor, n (%) Yes** |  | 2,454<br>(19.1) | 115,696<br>(24.2) | 118,150<br>(24) | 768<br>(23.1) | 32861<br>(27.6) | 33629<br>(27.4) |
| **Duration 2nd Stage, in minutes mean (SD)** |  | 44.5<br>(39.3) | 31.9<br>(37.8) | 32.2<br>(37.8) | 45.6<br>(38.0) | 30.7<br>(33.9) | 31.1<br>(34) |
| **Episiotomy, n (%)** | No | 10,635<br>(83) | 370,289<br>(77) | 380,924<br>(77) | 2746<br>(82.6) | 94565<br>(79.3) | 97311<br>(79.4) |
|  | Mediolat<br>eral | 2176<br>(16.9) | 107,467<br>(22) | 109,643<br>(22) | 568<br>(17.1) | 24487<br>(20.5) | 25055<br>(20.4) |
|  | Median | 48<br>(0.4) | 1119<br>(0.3) | 1167<br>(0.2) | 9<br>(8.3) | 146<br>(0.1) | 155<br>(0.12) |
| **Fetal sex, n (%)** | Girl | 5963<br>(46.4) | 237,044(4<br>9.5) | 243,007<br>(49) | 1540<br>(46.3) | 58838<br>(49.4) | 60378<br>(49.3) |
|  | Boy | 6896<br>(53.6) | 241,831<br>(50.5) | 248727<br>(51) | 1783<br>(53.7) | 60360<br>(50.6) | 62143<br>(50.7) |
| **Previous cesarean delivery, n (%)** | Yes | 626<br>(5.0) | 17,091<br>(3.5) | 17717<br>(3.6) | 160<br>(4.8) | 3918<br>(3.3) | 4078<br>(3.3) |
| **Mode of Delivery, n (%)** | Spontaneous | 11,331<br>(88) | 435,449<br>(91) | 446,780<br>(91) | 2913<br>(87.7) | 108,888<br>(91.4) | 111,801<br>(91.3) |
|  | Operative vaginal | 1,528<br>(12) | 43426<br>(9) | 44954<br>(9) | 410<br>(12.3) | 10,310<br>(8.6) | 10,720<br>(8.7) |
| **Epidural Analgesia, n (%)** | Yes | 2721<br>(21.2) | 99812<br>(20.8) | 102533<br>(20.9) | 818<br>(24.6) | 27212<br>(22.8) | 28030<br>(22.9) |
| **Parity, n (%)** | Nulliparous | 8,362<br>(65) | 201,073<br>(42) | 209,435<br>(42) | 2231<br>(67.1) | 50649<br>(42.5) | 52880<br>(43.2) |
|  | Primiparous | 3,833<br>(30) | 177,493<br>(37) | 181326<br>(37) | 931<br>(28) | 43798<br>(37) | 44,729<br>(36.5) |
|  | Multiparous | 664<br>(5) | 100309<br>(21) | 100973<br>(20) | 161<br>(4.8) | 24751<br>(20.8) | 24912<br>(20.3) |
| **Fetal distress n (%)** | Yes | 520<br>(4) | 22,255<br>(4.6) | 22775<br>(4.63) | 236<br>(7.1) | 9029<br>(7.5) | 9265<br>(7.5) |
| **Pregnancy diabetes n (%)** | Yes | 164<br>(1.28) | 7,593<br>(1.6) | 7757<br>(1.5) | 48<br>(1.4) | 2340<br>(1.9) | 2388<br>(1.9) |
| **Pregnancy Hypertension n (%)** | Yes | 277<br>(2.1) | 11331<br>(2.3) | 11608<br>(2.3) | 97<br>(2.9) | 3068<br>(2.5) | 3165<br>(2.5) |
| **Macrosomia ≥ 4000 grams, n (%)** | Yes | 2,853<br>(22.2) | 65,465<br>(13.7) | 68318<br>(13.9) | 689<br>(20.7) | 16660<br>(14) | 17349<br>(14.2) |
| **Prolonged labor, n (%)** | Yes | 548<br>(4.2) | 16941<br>(3.5) | 17489<br>(3.5) | 167<br>(5.1) | 5058<br>(4.2) | 5225<br>(4.2) |

### Model Performance

The MEGB model with macrosomia only (Model II) showed consistent performance across development and validation datasets, with AUCs of 0.729 and 0.727, respectively (95% CI: 0.720–0.735, Table 2A). Sensitivity and specificity were 0.668 and 0.664. Calibration was adequate (slope = 0.905, intercept = 0.003, Brier score = 0.026, Fig 3A). Hyperparameter tuning identified optimal settings of 250 trees, depth 5, and learning rate 0.05.

**Table 2A.** Internal and temporal validation measures of the OASIS predictive model with macrosomia only (Model II).

| **MODEL II : INCLUDING MACROSOMIA ONLY** |  |  |  |  |  |
| --- | --- | --- | --- | --- | --- |
| **Methods** | **Validation type** | **AUC and 95% CI** | **Intercept** | **Slope** | **Brier score** |
| **MEGB** | **Internal** | 0.7293 [0.725 – 0.733] | 0.002 | 1.06 | 0.025 |
|  | **External** | 0.7274 [0.719 – 0.735] | 0.003 | 0.905 | 0.026 |
| **ME-LASSO** | **Internal** | 0.7196 [0.715 – 0.723] | 0.000 | 1.014 | 0.025 |
|  | **External** | 0.7186 [0.710 – 0.727] | 0.001 | 0.997 | 0.026 |
| **Stepwise logistic regression** | **Internal** | 0.7182 [0.713 – 0.722] | 0.000 | 1.005 | 0.025 |
|  | **External** | 0.7144 [0.706 – 0.722] | 0.001 | 0.992 | 0.026 |

Variable importance analysis identified parity as the most influential predictor (22.7%), followed by second-stage duration (19.1%), episiotomy (17.7%), and macrosomia (13.5%) (Fig 2A).

**Fig 2A.**
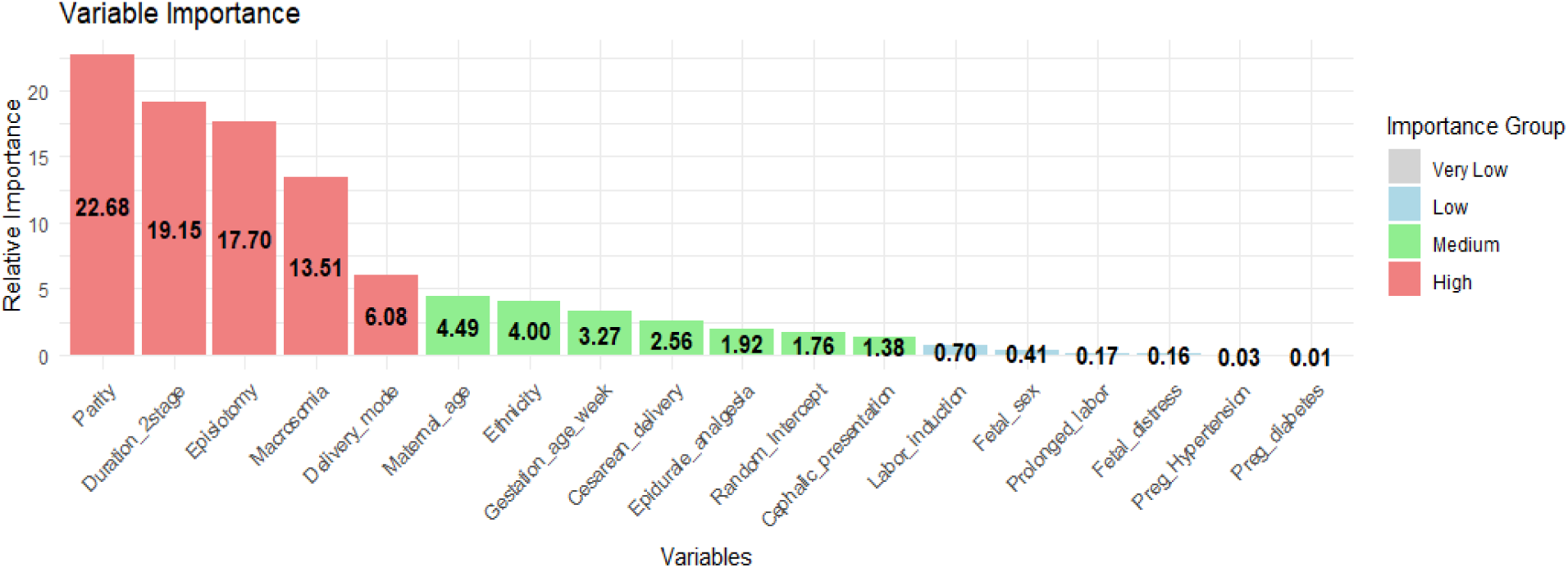
Variable importance from the MEGB model (Model II: macrosomia only). Relative importance of predictors for obstetric anal sphincter injury (OASIS) estimated using gradient boosting within the mixed-effects machine learning framework, with a random intercept for hospital-level variation. The most influential predictors were parity, second-stage duration, episiotomy, macrosomia, delivery mode, maternal age, and ethnicity.

Slightly better results were observed for the model including exact birth weight (Model I); details are provided in (Table 3B, Fig 2B and S1 Fig).

**Fig 2B.**
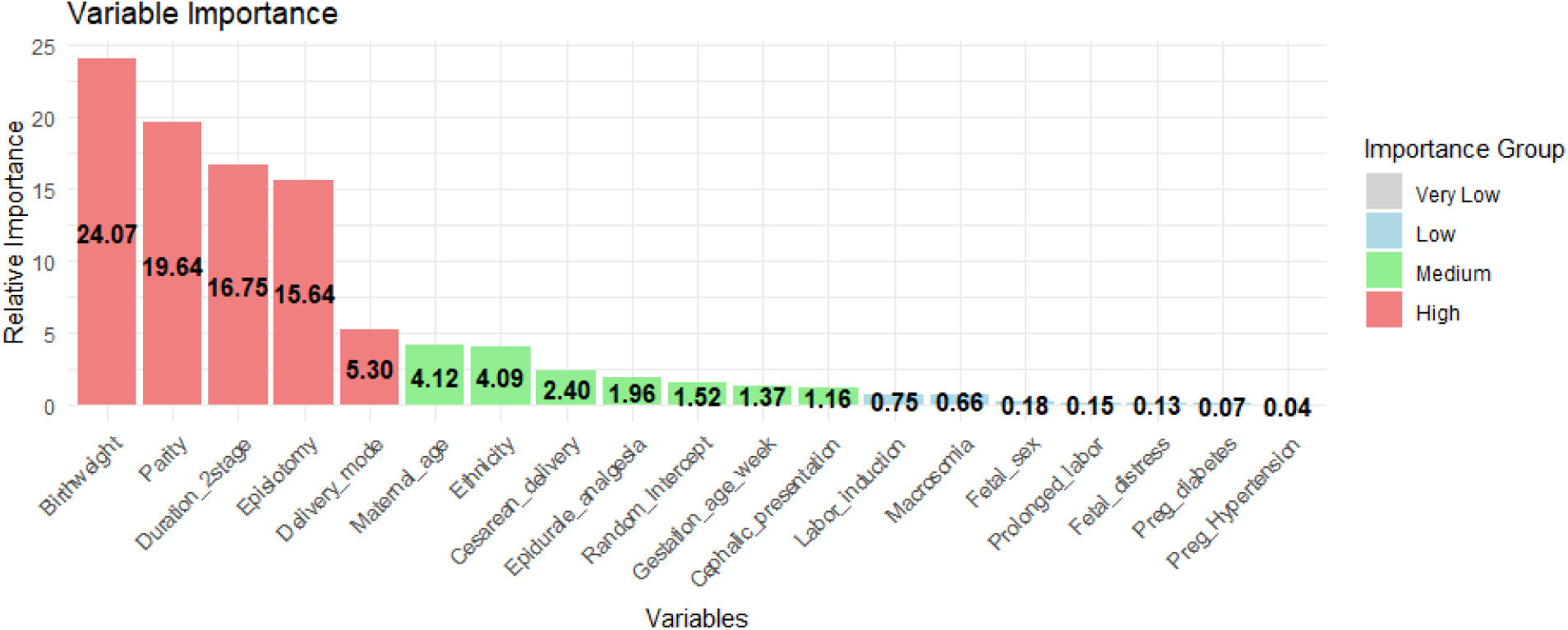
Variable importance from the MEGB model (Model I: Including exact birth weight). Relative importance of predictors for obstetric anal sphincter injury (OASIS) estimated using the GBM algorithm within the mixed-effects machine learning (MEML) framework. Random intercept reflects hospital-level variation. Key predictors with high or medium importance included exact birth weight, parity, second-stage duration, episiotomy, delivery mode, maternal age and ethnicity.

**Table 2B.** Internal and temporal validation measures of the OASIS predictive model including exact birth weight (Model I).

| **MODEL I : INCLUDING EXACT BIRTH WEIGHT** |  |  |  |  |  |
| --- | --- | --- | --- | --- | --- |
| **Methods** | **Validation type** | **AUC and 95% CI** | **Intercept** | **Slope** | **Brier score** |
| **MEGB** | **Internal** | 0.7399 [0.735 – 0.744] | 0.004 | 1.134 | 0.025 |
|  | **External** | 0.7394 [0.731 – 0.747] | 0.002 | 0.929 | 0.026 |
| **ME-LASSO** | **Internal** | 0.7330 [0.728 – 0.737] | 0.000 | 0.998 | 0.025 |
|  | **External** | 0.7318 [0.723 – 0.740] | 0.000 | 1.024 | 0.026 |
| **Stepwise logistic regression** | **Internal** | 0.7315 [0.727 – 0.735] | 0.000 | 0.996 | 0.025 |
|  | **External** | 0.7283 [0.719 – 0.736] | 0.001 | 0.985 | 0.026 |

The ME-LASSO Model II achieved an internal AUC of 0.720 and an external AUC of 0.719 (95% CI: 0.710–0.727, Table 3A), with sensitivity of 0.66 and specificity of 0.65. Isotonic calibration yielded good agreement between predicted and observed risks (slope = 0.997, intercept = 0.001, Brier score = 0.026, Fig 3B). All candidate predictors were retained except pregnancy hypertension (Table 2A). The model including exact birth weight (Model I) yielded slightly better performance (p<0.001), as shown in (Table 2B, Table 3B and S2 Fig).

**Fig 3A.**
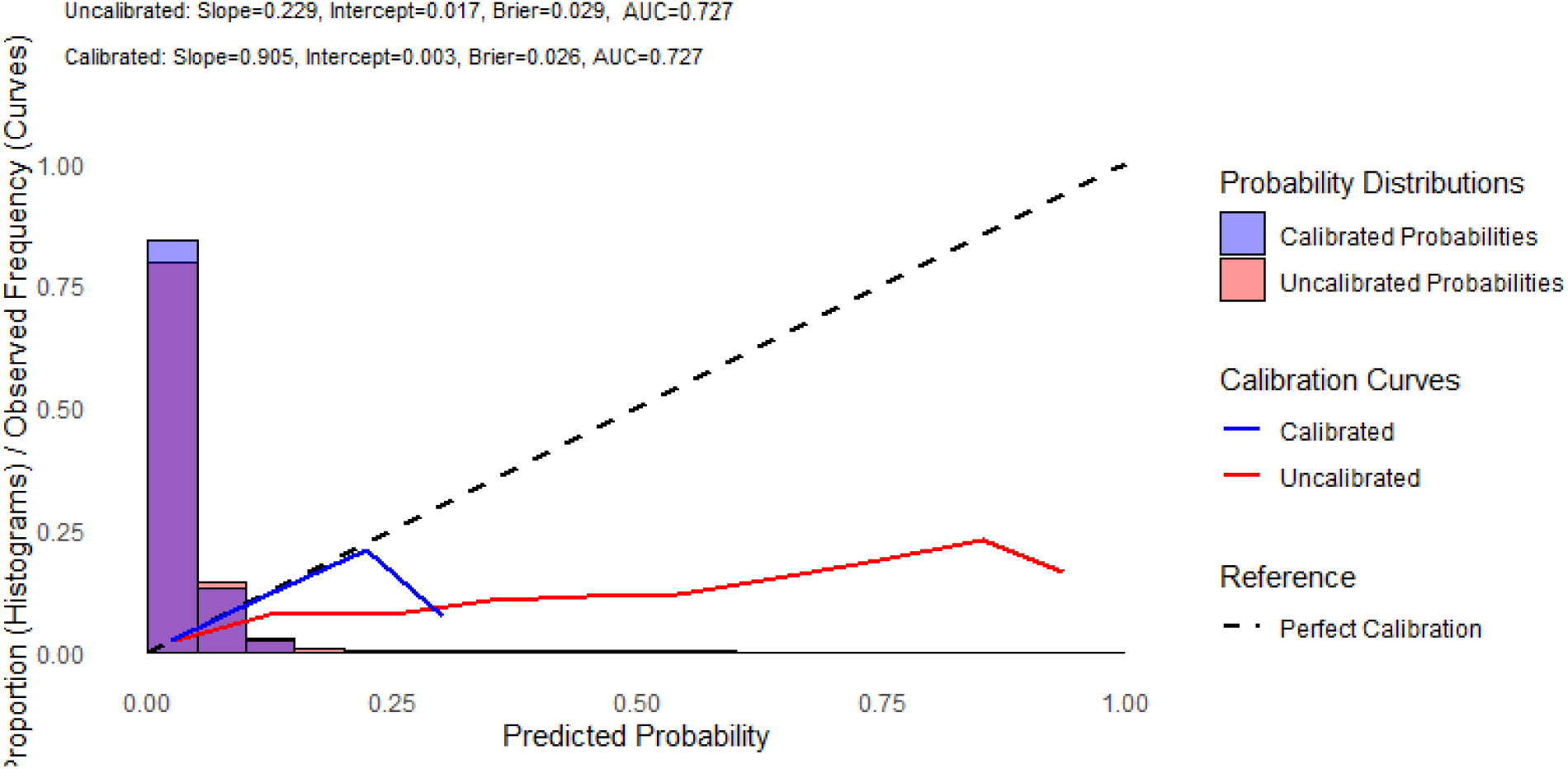
Calibration plot for external validation of OASIS using MEGB (Model II: macrosomia only).

**Fig 3B.**
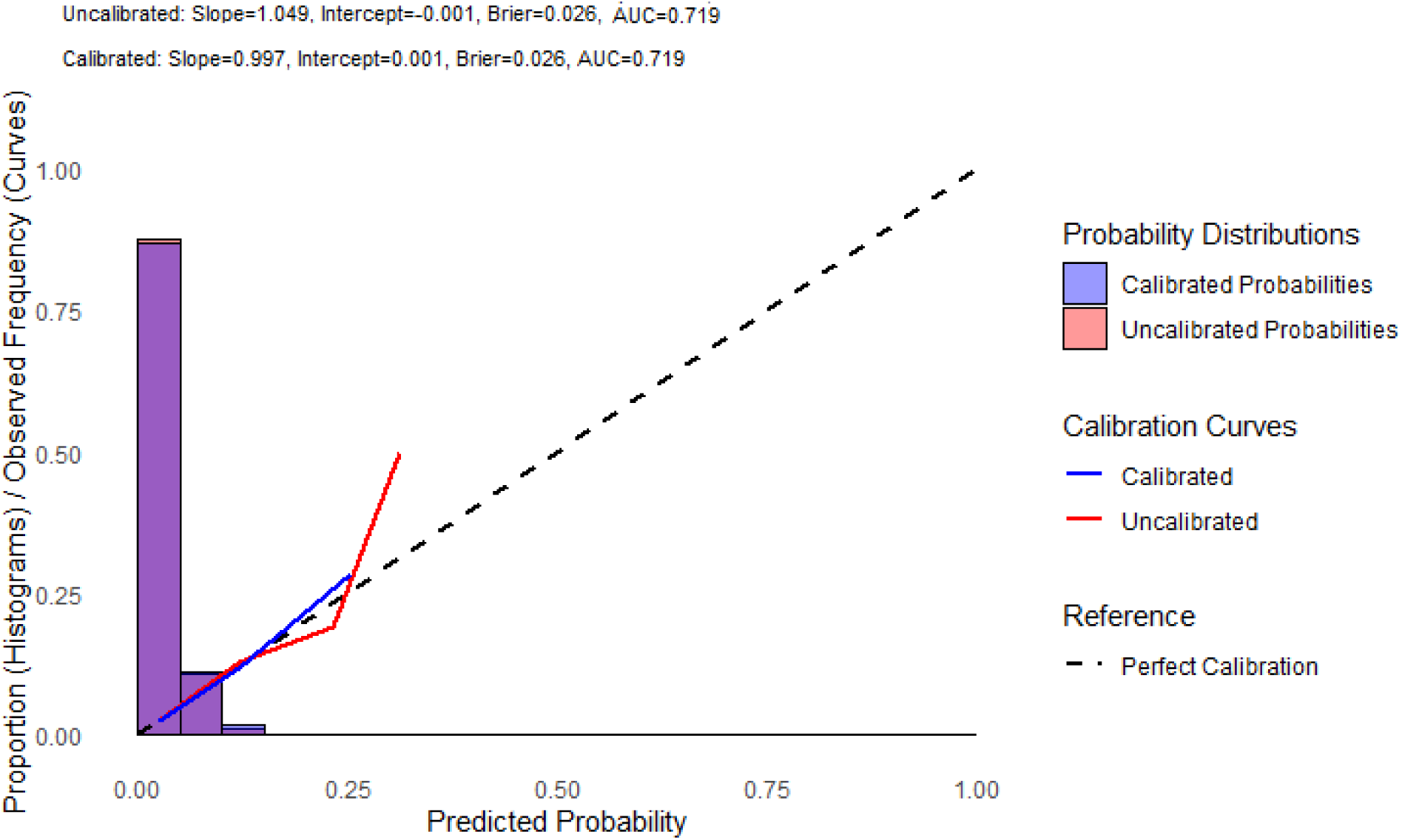
Calibration plot for external validation of OASIS using ME-LASSO (Model II: macrosomia only).

**Table 3A.**
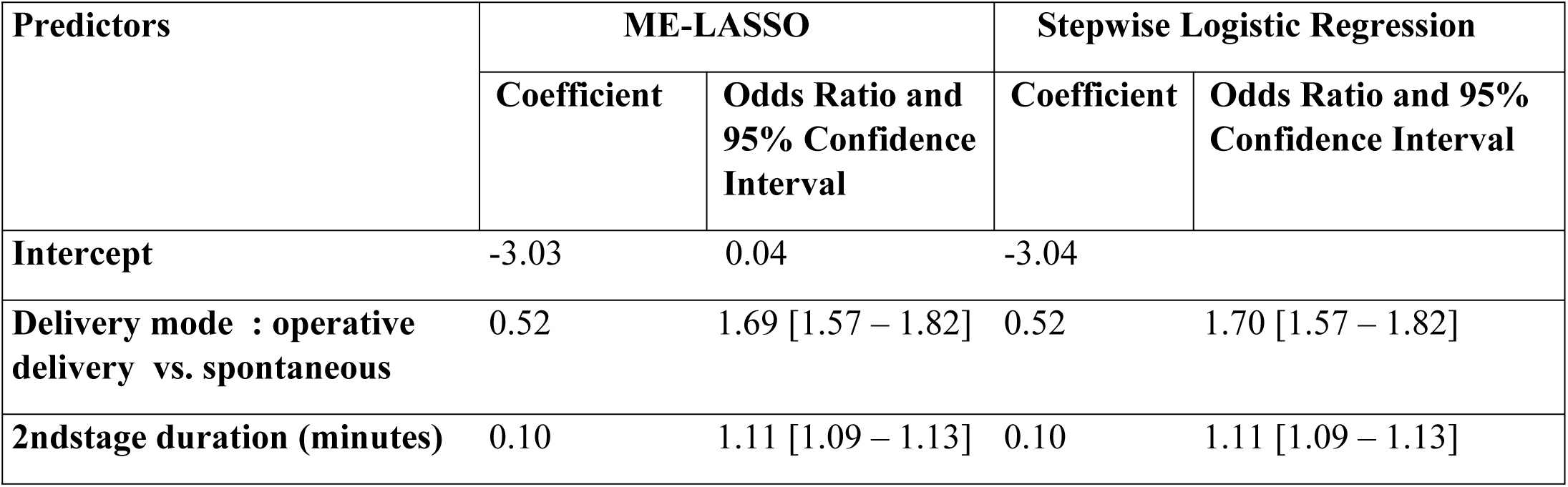
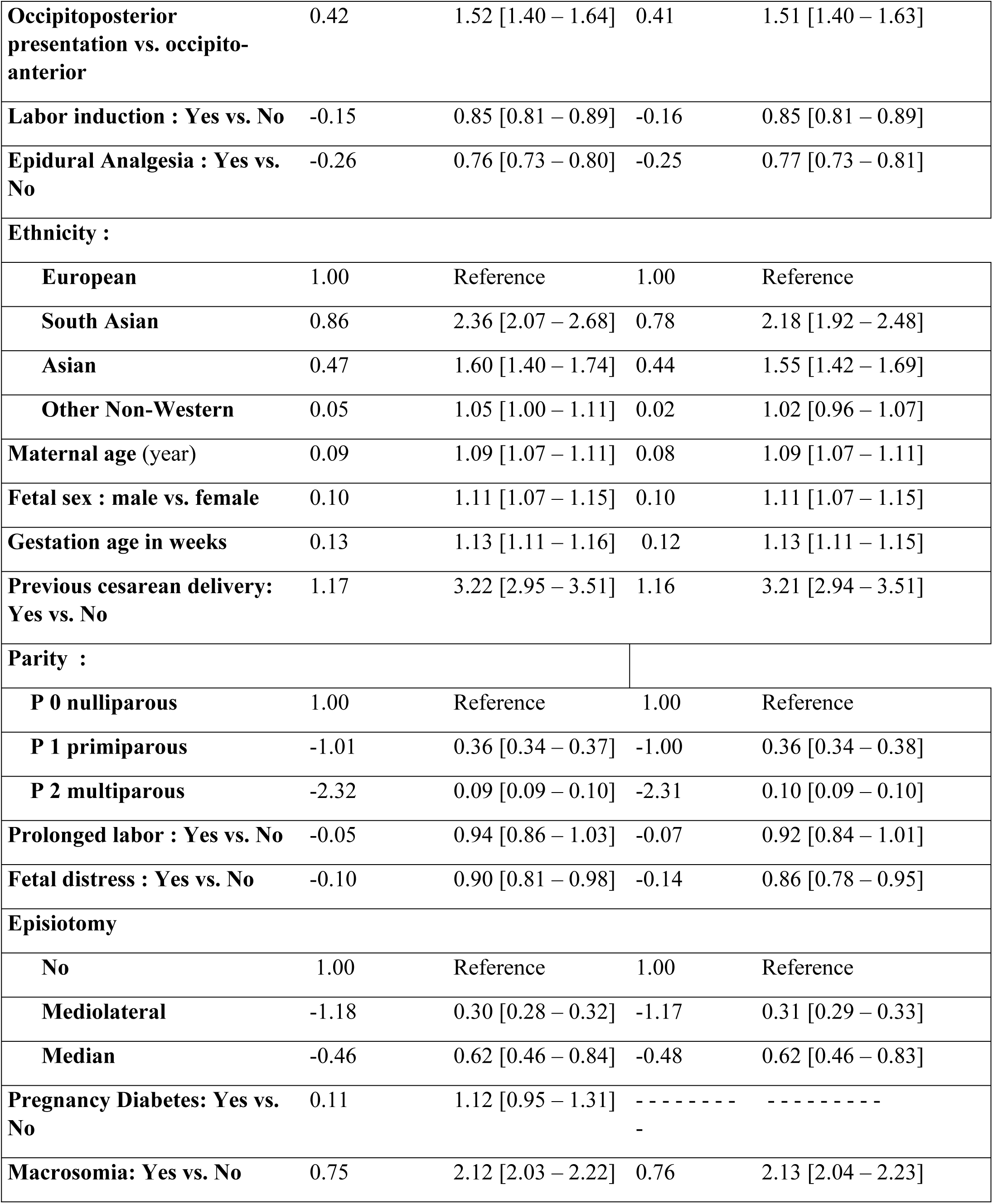
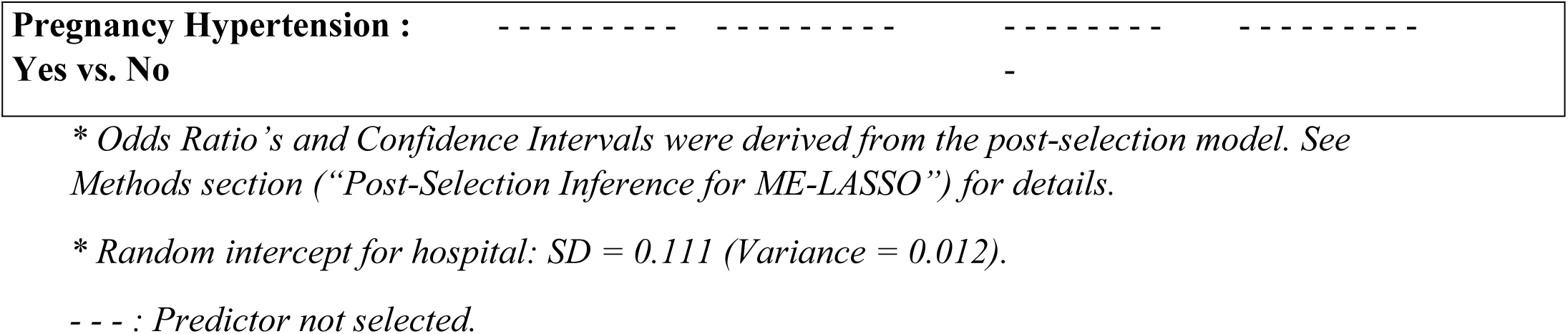
Coefficients and odds ratios for OASIS prediction from ME-LASSO and stepwise logistic regression (Model II: macrosomia only).

**Table 3B.** Coefficients and odds ratios for OASIS prediction from ME-LASSO and stepwise logistic regression (Model I: including exact birth weight).

| Predictors | ME-LASSO |  | Stepwise Logistic Regression |  |
| --- | --- | --- | --- | --- |
|  | Coefficient | Odds Ratio and 95% Confidence Interval | Coefficient | Odds Ratio and 95% Confidence Interval |
| Intercept | -2.88 | 0.05 [0.05 – 0.06] | -2.89 | 0.06 [0.05 – 0.05] |
| Delivery mode: Operative | 0.53 | 1.69 [1.58 – 1.82] | 0.57 | 1.77 [1.62 – 1.93] |
| Birth weight (grams) | 0.46 | 1.59 [1.54 – 1.63] | 0.48 | 1.62 [1.58 – 1.65] |
| 2nd stage duration (minutes) | 0.09 | 1.09 [1.07 – 1.11] | 0.09 | 1.09 [1.07 – 1.11] |
| Occipitoposterior presentation vs. occipitoanterior | 0.43 | 1.53 [1.41 – 1.65] | 0.46 | 1.59 [1.45 – 1.75] |
| Labor induction : Yes vs. No | -0.14 | 0.87 [0.83 – 0.91] | -0.15 | 0.85 [0.81 – 0.90] |
| Epidural Analgesia : Yes vs. No | -0.27 | 0.76 [0.72 – 0.80] | -0.25 | 0.77 [0.73 – 0.82] |
| Ethnicity : |  |  |  |  |
| European |  | Reference |  | Reference |
| South Asian | 1.08 | 2.94 [2.58 – 3.34] | 0.98 | 2.67 [2.28 – 3.13] |
| Asian | 0.56 | 1.76 [1.61 – 1.92] | 0.60 | 1.82 [1.64 – 2.02] |
| Other Non-Western | 0.11 | 1.11 [1.06 – 1.17] | 0.07 | 1.07 [1.00 – 1.14] |
| Maternal age (year) | 0.1 | 1.10 [1.08 – 1.12] | 0.08 | 1.09 [1.06 – 1.11] |
| Fetal sex : male vs. female | 0.03 | 1.03 [0.99 – 1.07] | ----- | ----- |
| Gestation age in weeks | 0.01 | 1.01 [0.98 – 1.03] | ----- | ----- |
| **Previous cesarean delivery: Yes vs. No** | 1.21 | 3.36 [3.07 – 3.67] | 1.20 | 3.34 [3.00 – 3.72] |
| **Parity :** |  |  |  |  |
| **P 0 nulliparous** |  | Reference |  | Reference |
| **P 1 primiparous** | -1.12 | 0.32 [0.31 – 0.34] | -1.11 | 0.32 [0.30 – 0.34] |
| **P 2 multiparous** | -2.46 | 0.08 [0.07 – 0.09] | -2.44 | 0.08 [0.07 – 0.09] |
| **Prolonged labor : Yes vs. No** | -0.09 | 0.91 [0.83 – 1.00] | -0.11 | 0.90 [0.80 – 1.00] |
| **Fetal distress : Yes vs. No** | -0.04 | 0.96 [0.87 – 1.05] | -0.09 | 0.91 [0.81 – 1.02] |
| **Episiotomy:** |  |  |  |  |
| **No** | 1.00 | Reference | 1.00 | Reference |
| **Mediolateral** | -1.2 | 0.30 [0.28 – 0.31] | -1.18 | 0.30 [0.28 – 0.33] |
| **Median** | -0.48 | 0.61 [0.46 – 0.83] | -0.67 | 0.50 [0.34 – 0.75] |
| **Pregnancy Hypertension : Yes vs. No** | 0.03 | 1.03 [0.91 – 1.17] | ----- | ----- |
| **Pregnancy Diabetes: Yes vs. No** | ----- | ----- | ----- | ----- |
| **Macrosomia: Yes vs. No** | 0.03 | 1.03 [0.97 – 1.09] | ----- | ----- |

\* Odds Ratio 's and Confidence Intervals for ME-LASSO were derived from the post-selection model. See Methods section ("Post-Selection Inference for ME-LASSO") for details.
\* Random intercept for hospital: SD = 0.110 (Variance = 0.012).
- - - : Predictor not selected.

In the backward stepwise logistic regression model with macrosomia only (Model II), the internal AUC was 0.718 and a temporal AUC of 0.714 (95% CI: 0.706–0.722, Table 3A), with sensitivity of 0.66 and specificity of 0.65. All the top 10 ranked variables by importance in the MEGB model were selected in the logistic regression model. Predictor variables pregnancy hypertension and diabetes were not retained (Table 2A). Calibration was excellent (slope = 0.992, intercept = 0.001, Brier score = 0.026, Fig 3C). Results for the model including birth weight (Model I) were similar and slightly improved as reported in (Table 2B, Table 3B and S3 Fig).

**Fig 3C.**
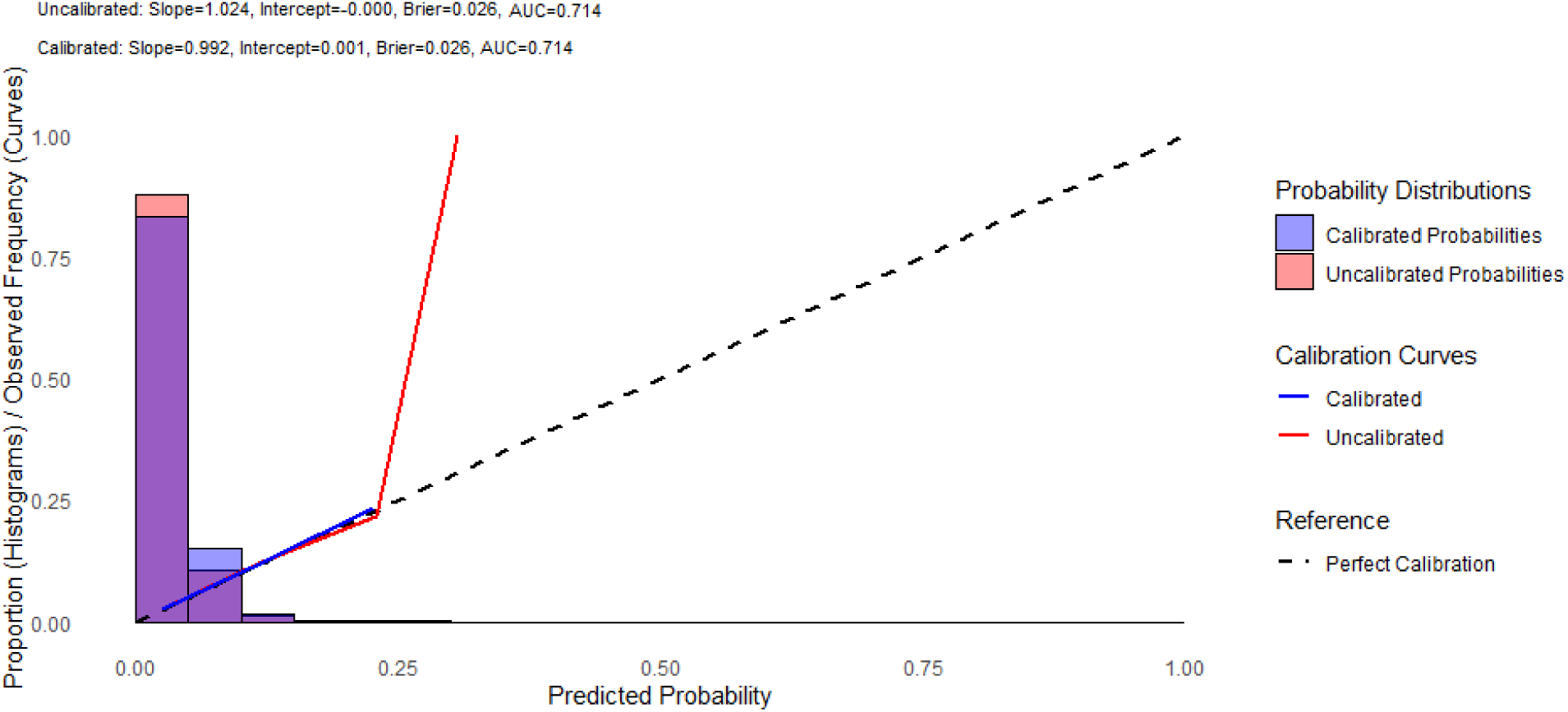
Calibration plot for external validation of OASIS using stepwise logistic regression (Model II: macrosomia only).

## Discussion

### Principal findings

Both mixed-effects models outperformed stepwise logistic regression for OASIS risk prediction, with MEGB statistically significantly outperforming ME-LASSO, although the absolute difference was small. In temporal validation, MEGB achieved the highest discriminative accuracy (AUC 0.727, 95% CI: 0.719–0.735), outperforming ME-LASSO (AUC 0.719, 95% CI: 0.710–0.727) and stepwise logistic regression (AUC 0.714, 95% CI: 0.706–0.722). These results correspond to Model II using macrosomia only. Including birth weight yielded only marginal improvements (AUC increase 0.01–0.02), as macrosomia served as an effective proxy. Performance was consistent between internal and temporal validation, with only marginally stronger performance in internal validation, indicating robust generalization across time. While uncalibrated MEGB initially exhibited poorer calibration, isotonic recalibration achieved excellent alignment (slope ≈1.0), comparable to ME-LASSO and stepwise logistic regression, highlighting the importance of recalibration for clinical deployment of machine learning models.

Across all three methods, MEGB, ME-LASSO, and stepwise logistic regression, parity (with nulliparous women showing the highest risk), longer second-stage duration, operative delivery mode, and mediolateral episiotomy (associated with lower risk) consistently emerged as the strongest predictors of OASIS.

### Results of the study in context

Mixed-effects methods such as MEGB and ME-LASSO are particularly valuable when patients are clustered within hospitals or regions: by adjusting for between-hospital differences in patient populations and clinical practices, they yield more generalizable individual predictions. Such methods have been applied beyond obstetrics, including in cardiology, cancer research, neonatology, and medical imaging [26–28].

Our predictors align with the OASIS literature: multiparity and mediolateral episiotomy were associated with a significantly lower risk [4,8], while nulliparity, operative vaginal delivery, advanced maternal age, prolonged second stage, (South) Asian ethnicity, and prior cesarean were associated with higher risk [1,3,4,8]. High birth weight (or macrosomia as its proxy) consistently ranked among the top predictors, reinforcing its well-established association with increased risk of sphincter injury [1,2,4].

Epidural analgesia and labor induction were associated with lower risk, consistent with some prior findings [29] though differing from more recent evidence [30]. Pregnancy hypertension, diabetes, and male fetal sex lacked predictive value in adjusted models, consistent with prior evidence [31].

A recent systematic review on prediction models for OASIS reported discrimination of prior OASIS prediction models ranging from 0.64 to 0.83. The authors found that all models had methodological shortcomings, including small or poorly generalizable cohorts and risk of bias, and that none had undergone geographical external validation, so even the higher discrimination values may be optimistic [10]. Our temporally validated AUC of 0.727, obtained in a nationwide cohort with adjustment for hospital-level clustering, offers a more realistic and transportable estimate of discrimination. Rather than identifying novel determinants, our approach improves individualized prediction by modeling nonlinear effects, accounting for hospital-level heterogeneity, and achieving robust discrimination with comparable calibration.

### Clinical Implications

The MEGB Model II achieved an AUC of 0.72 in clinical risk prediction: meaning it correctly ranks an OASIS patient as higher risk than a non-OASIS patient 72% of the time. The consistent prominence of well-established risk factors across all models supports communication on risks and targeted antenatal and intrapartum interventions for high-risk patients.

Advancements in antenatal weight estimation by vigorous ultrasound examination, combined with our finding that macrosomia effectively substitutes for birth weight, support using a simple dichotomous macrosomia predictor variable, rather than exact fetal weight estimates.

Furthermore, using a model with macrosomia enhances the clinical applicability of the model in daily practice as the use of an uncertain factor of major importance is largely avoided.

Finally, while MEGB showed the highest discrimination, prospectively validated models may support individual risk assessment and help target preventive care.

### Research Implications

Though significant, the limited incremental gain of MEGB compared with ME-LASSO likely reflects constraints inherent to registry-based predictors. Important predictors such as maternal BMI, smoking status, prior OASIS history, and intrapartum factors including manual perineal support were not available in our dataset and may explain residual variation in OASIS prediction. Future research should evaluate whether MEGB’s advantages become more pronounced when incorporating such clinically relevant variables beyond those captured in routine registry data.

Further work should externally validate our model in diverse populations and examine implementation feasibility. Since we relied on logistic regression for interpreting predictors, future studies should apply model-agnostic explainability techniques, such as SHapley Additive exPlanations (SHAP) values, to MEGB to clarify how each individual feature contributed to the predictions and enhance transparency [32].

Additionally, prospective validation should assess whether our macrosomia-based model (Model II), using antenatal ultrasound estimation of macrosomia, can effectively stratify OASIS risk before delivery. This would translate our finding that dichotomous macrosomia substitutes for exact birth weight into a clinically implementable antenatal risk assessment tool.

Together, these directions may improve the performance, interpretability, and clinical utility of OASIS risk models, supporting individualized counseling and targeted preventive care in obstetrics.

### Strengths and limitations

A major strength of this study is the use of a large, nationwide perinatal registry including 614,255 vaginal deliveries, offering strong statistical power and broad population coverage. The multicenter design, spanning all Dutch healthcare regions, enabled the application of mixed-effects frameworks incorporating both fixed and random effects, alongside advanced machine learning (MEGB, ME-LASSO) and stepwise logistic regression approaches. MEGB captures non-linear relationships without pre-specification, and multilevel imputation appropriately handled missing data within the hierarchical structure.

To our knowledge, this is among the first study to apply MEGB and ME-LASSO for OASIS prediction while accounting for region-level heterogeneity. Key steps, including temporal validation, multilevel imputation, and post hoc isotonic calibration, were conducted in accordance with TRIPOD+AI guidelines to enhance model reliability and generalizability. Both MEGB and ME-LASSO addressed key challenges in prediction modeling: accounting for clustering via hierarchical modeling and automating variable selection to improve interpretability.

Unlike earlier OASIS models, often limited by single-center data or absence of mixed-effects structures, our study addresses institutional heterogeneity and combines traditional and modern methods. MEGB showed stable performance across development and validation datasets, indicating reduced overfitting and enhanced generalizability.

However, several limitations exist. MEGB poses challenges in interpretability, complexity, and computational demands, while ME-LASSO may exclude clinically relevant predictors. Both are vulnerable to algorithmic bias if subpopulations are underrepresented. Additionally, interpretation of risk factors relied on the regression model’s outputs, not MEGB directly.

Turning to the data, macrosomia was defined from recorded birth weight; antenatal use would rely on estimated fetal weight (antenatally estimable macrosomia), an approach we did not validate here. Several predictors, such as episiotomy, prolonged second stage, and operative delivery, are intrapartum and not known antenatally; although they can be varied to explore delivery scenarios, this yields conditional rather than absolute risk estimates. The registry also lacked key predictors known to be associated with OASIS risk, such as maternal BMI and previous OASIS, possibly reducing predictive precision. Although temporally validated, generalizability to other healthcare systems remains uncertain and requires external validation.

Baseline characteristics and OASIS incidence were comparable between 2020 and 2016-2019 (Table 1), minimizing concerns about possible COVID-19-related practice changes. Nonetheless, this study demonstrates how integrating machine learning and mixed-effects modeling can advance scalable, data-driven OASIS risk prediction in clinical care. An interactive online calculator based on MEGB model II is available for illustrative purposes: https://kikresearch-apps.amsterdamumc.org/oasis-megb/. This online calculator requires clinicians to manually enter patient-specific predictors (e.g., parity, second-stage duration, episiotomy type, macrosomia status). No specialized technical skills are needed beyond routine obstetric knowledge. For use of the prediction model outside the Netherlands, or in hospitals not represented in the development data, the model defaults to fixed-effect predictions only, as the hospital-specific random effect cannot be estimated. The online calculator applies input validation to the entered predictor values and requires all predictors to be provided before a prediction is generated, so imputation of missing values at the point of care is not supported in the current implementation. Given the low proportion of missing values, the influence of the imputation strategy on reported performance estimates is expected to be minimal, though reported metrics reflect a batch-imputation scenario, consistent with anticipated use on periodically extracted registry data, rather than real-time imputation for individual patients. In a future implementation study, imputation models for the two affected variables could be re-estimated on the full cohort and deployed alongside the final model, excluding the outcome since OASI is unknown at the time of prediction; their transportability to external cohorts would assume broadly similar missingness mechanisms and case-mix, which cannot be verified a priori. This applies mainly to the regression-based models, as gradient boosting handles missing values natively; data were imputed before model fitting to keep the comparison between approaches on equal footing.

## Conclusions

Mixed-effects gradient boosting (MEGB), which accounts for hospital-level heterogeneity, demonstrated higher discriminative performance and comparable calibration compared to Backward Stepwise Logistic Regression, with MEGB performing slightly better than ME-LASSO for OASIS prediction, with consistent performance across development and validation cohorts. Importantly, models using macrosomia as a proxy for exact birth weight performed nearly as well, potentially supporting their use for antenatal risk assessment if macrosomia is estimated prenatally. Integrating these advanced ML models into clinical tools may enhance individualized prediction and help target preventive care.

## Declaration of generative AI and AI-assisted technologies in the manuscript preparation process

During the preparation of this work the author(s) used Claude, ChatGPT, and Perplexity to assist with language editing. The author(s) reviewed and edited the output as needed and take full responsibility for the content of the published article.

## Data Availability

The data underlying this study were obtained from the Dutch Perinatal Registry (Perined). The authors do not have permission to share these data publicly, as they are subject to third-party governance and privacy restrictions under Perined's data-access regulations. Data are available to researchers who meet the criteria for access to confidential data upon reasonable request to Perined (https://www.perined.nl), subject to Perined's application and approval procedures. The authors did not have special access privileges that others would not have. Analysis code is available at https://github.com/achahid/OASI-prediction-model."

https://github.com/achahid/OASI-prediction-model

## Supporting information caption

**S1 Fig. Calibration plot for external validation of OASIS using MEGB (Model I: including exact birth weight).**

**S2 Fig. Calibration plot for external validation of OASIS using ME-LASSO (Model I: including exact birth weight).**

**S3 Fig. Calibration plot for external validation of OASIS using stepwise logistic regression (Model I: including exact birth weight).**

**S4 Checklist. STROBE checklist.**

**S5 Checklist. TRIPOD+AI checklist.**

